# Outbreak of SARS-CoV-2 in a prison: Low effectiveness of a single dose of the adenovirus vector ChAdOx1 vaccine in recently vaccinated inmates

**DOI:** 10.1101/2021.08.03.21258337

**Authors:** A Marco, N Teixidó, RA Guerrero, L Puig, X Rué, A Cobo, I Barnés, E Turu

## Abstract

**Introduction:** To analyse the effectiveness of a dose of adenovirus vector ChAdOx1 vaccine (AVChOx1) in an outbreak of SARS-CoV-2 detected in a prison.

**Methods:** Observational study carried out at Brians-1 Prison, Barcelona. After detecting a case of infection, rt-PCR was administered to all prisoners (some of whom had been vaccinated 21-23 days previously with a dose of AVChOx1) and to staff. Infection rates in vaccinated and unvaccinated populations were calculated, as was vaccine effectiveness.

**Results:** One hundred and eighty-four asymptomatic prisoners (50.3% vaccinated) and 33 staff were screened. Forty-eight (25.9%) infections by the SCV-B.1.17 variant were recorded in prisoners and none in staff. Infection rates were higher in younger prisoners, immigrants, and those admitted ≥7 days previously. In all, 22.6% of vaccinated subjects were infected vs. 29.3% of unvaccinated. Vaccine effectiveness was 23%. Only 6.2 cases would have been prevented by vaccinating the unvaccinated individuals. At seven days, the rt-PCR was negative in 46.2% of vaccinated subjects vs. 13.6% of unvaccinated (p = 0.02).

**Discussion:** In a prison outbreak, a dose of AVChAdOx1 administered 21-23 days earlier did not significantly prevent the occurrence of infections, but did reduce the duration of rt-PCR positivity. Maintaining post-vaccination preventive measures is essential.

## Introduction

In prisons, interpersonal contact and the high density of occupation have favored the transmission of SARS-CoV-2. In fact, only 27 days after the first case detected in a Catalan prison, the first outbreak occurred, affecting 40 inmates, two health workers, and four prison guards^1^.

Many guides have been published by national and international agencies regarding the management and control of SARS-CoV-2 infection in prisons. In general, they coincide in terms of the measures and procedures to be used. However, control will only be fully effective when there is high vaccination coverage (this is the case outside prison as well). Currently, there are more than 200 vaccines in development and more than 60 are in the clinical phase. In Spain, the use of four vaccines has been approved: two using messenger RNA technology (mRNA-1273 SARS-CoV-2, and BNT162b2 mRNA Covid-19) and two based on viral vectors (adenovirus vector ChAdOx1, and Ad26.COV2.S). As of May 10, more than 20 million doses had been administered: more than 14 million people with one dose, and 6.3 million (13.3% of the population) with the full regimen^2^. These vaccines generate immunity, but certain aspects remain unknown – such as the duration of this immunity, the effectiveness against new variants, and the effectiveness in avoiding transmission.

The aims of the present study are to describe an outbreak of SARS-CoV-2 detected in a prison, to assess the infection rate observed in vaccinated and unvaccinated prisoners, and to calculate the effectiveness of a dose of the adenovirus vector ChAdOx1 (AVChAdOx1) vaccine administered 21-23 days previously.

## Methods

Observational study of inmates in module 2 (M2) of Brians-1 Prison, Barcelona. A real-time reverse transcription polymerase chain reaction test (rt-PCR) with a sample of nasopharyngeal/oropharyngeal exudate was performed in an inmate of M2 inmate. Twenty-one days earlier, along with other inmates, this prisoner had received the first dose of AVChAdOx1 vaccine. The rt-PCR result was positive. The module housing 185 inmates was isolated and all were screened with rt-PCR, as well as staff members who had worked in the prison during the previous seven days. All the samples were analysed and sequenced in the laboratory at Bellvitge University Hospital.

The M2 was sequentially disinfected and divided into three independent zones: a) zone “A” for cases with a positive rt-PCR result; b) zone “B” for cases with a negative rt-PCR result but who previously lived with an inmate with a positive result (close contacts); and c) zone “C” for cases with a negative rt-PCR result who had not lived with infected persons. Only essential health workers and security workers were allowed to enter the M2.

Cases with negative rt-PCR were screened at seven days and again in all cases after 14 days of isolation. In the cases with a positive result, the isolation was maintained and weekly rt-PCR was scheduled, adding cycle threshold values in SARS-CoV-2 to rule out cases with residual rt-PCR.

Infection rates in vaccinated and non-vaccinated patients, attributable risk (AR) and vaccine effectiveness were calculated, estimating how many unvaccinated individuals would have avoided infection had they been vaccinated.

### Ethical considerations

This observational study presents the results of the rt-PCR used for screening in accordance with our clinical practice. The tests were not requested for research purposes. However, the patients were informed of the purpose of data collection and their written informed consent was requested. The study was evaluated and approved by the Clinical Research Ethics Committee of the Jordi Gol Foundation (CEI: 21/169-PCV).

## Results

After detection of the index case, 184 prisoners and 33 workers were screened within 24 hours (Figure 1). All inmates were male and asymptomatic. Twenty-nine (15.7%) had resided in the M2 for <7 days and 156 (84.3%) for ≥7 days. Mean age was 38.6 years (range: 20-62); 29.7% were Spanish and five (2.7%) were HIV-infected.

**Figure 1.**
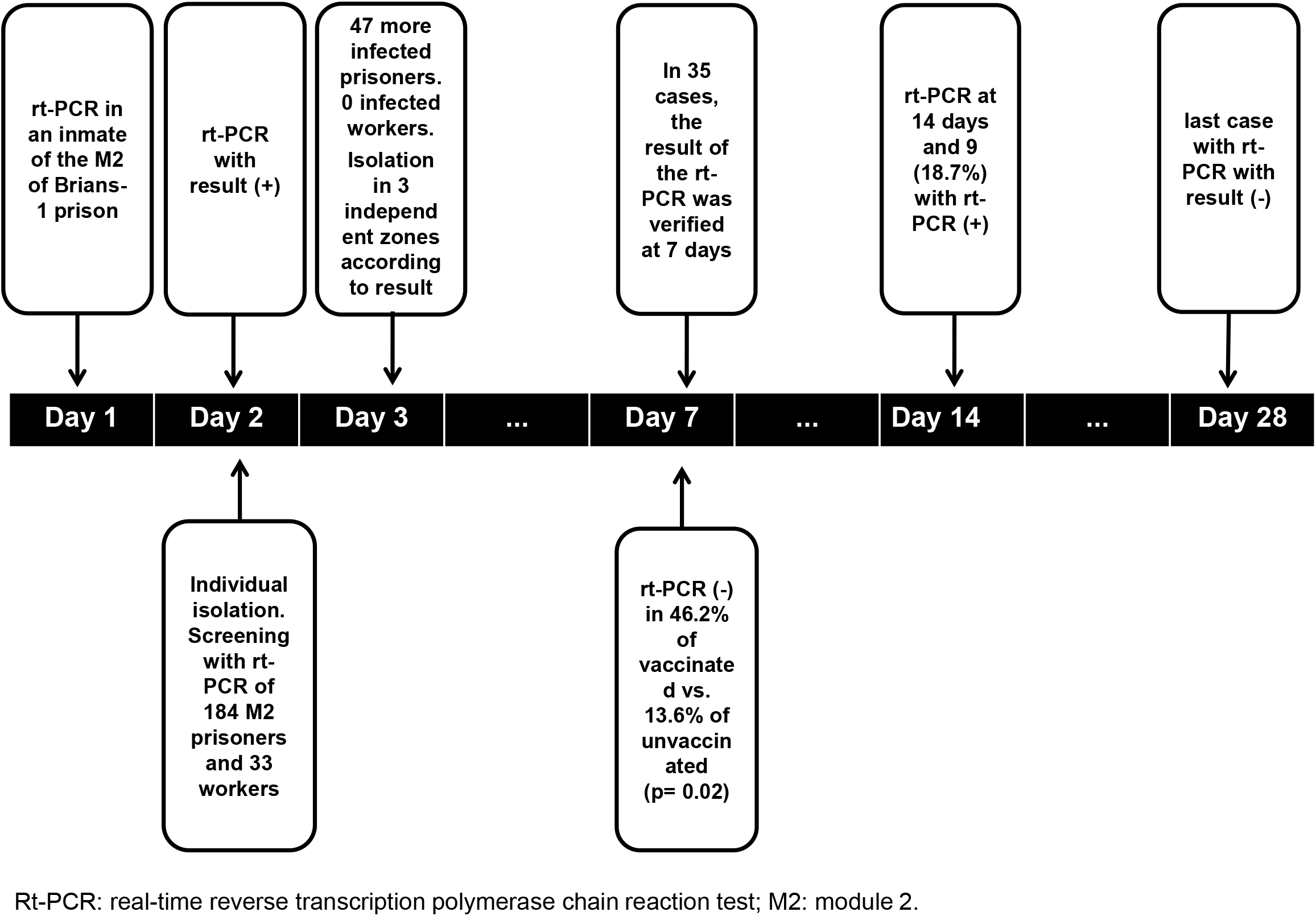
Measures taken and results of the screenings performed of the inmates of module 2 in Brians1 Prison, Barcelona from the detection of the first case of infection by the variant SARS-CoV-2 B.1.1.7 until the last case presented a negative rt-PCR.

Ninety-three (50.3%) had received a dose of AVChAdOx1 vaccine 21-23 days previously. Two (1.1%) who had presented COVID-19 in the last three months were not vaccinated, 45 (24.3%) refused vaccination and another 45 (24.3%) were recent admissions who had not yet been vaccinated. Rejection of vaccination was only associated with immigrant status (10.3% non-immigrant vs. 30% immigrant; p = 0.002).

The screening detected another 47 (25.9%) inmates infected with the SCV-B.1.17 variant, but no staff members. Infection was associated with: a) younger age (mean age was 37.8 years in infected subjects vs 41.3 years in non-infected; p = 0.01); b) length of stay in the M2 (one case <7 days vs 47 cases ≥7 days; p = 0.001); and c) immigrant status (31.5% in immigrants vs. 12.7% in non-immigrants; p = 0.003).

The rt-PCR was positive in 21 (22.6%) vaccinated subjects and in 27 (29.3%) unvaccinated subjects (p = 0.15). The AR was 6.8, and vaccine effectiveness was 23% (Table 1). According to these data, if the 27 unvaccinated infected subjects had been vaccinated, 6.2 cases would have been avoided.

**Table 1.** Incidence of infection by the variant SARS-CoV-2 B.1.1.7 in vaccinated and unvaccinated individuals, attributable risk and vaccine effectiveness of a single dose of adenovirus vector ChAdOx1 vaccine, administered 21-23 days previously, in an outbreak detected in Module 2 of Brians-1 Prison, Barcelona.

| **Infected/Vaccinated\***<br>**(Incidence/100)** | **Infected/Not Vaccinated**<br>**(Incidence/100)** | **Infected/Total**<br>**(Incidence/100)** | **AR** | **VE** |
| --- | --- | --- | --- | --- |
| 21/93<br>(22.6) | 27/92<br>(29.3) | 48/185<br>(25.9) | 6.8 | 23% |

(\*): Vaccinated with a single dose of ChAdOx1-S (recombinant) 21- 23 days previously.
AR: attributable risk
VE: vaccine effectiveness

All patients evolved satisfactorily. The result of the rt-PCR was checked at seven days in 35 cases, and was negative in 46.2% of vaccinated subjects vs. 13.6% of unvaccinated (p = 0.02). Nine patients (18.7%) still had positive rt-CRP at 14 days, and one up to 28 days later.

## Discussion

The effectiveness of a dose of AVChAdOx1 vaccine for preventing new infections was 23%. Only 6.2 cases would have been avoided if the unvaccinated subjects had been vaccinated. This low rate may be due to the fact that the vaccine is effective after 10-13 days, but does not achieve maximum protection until some weeks later^3^. In all inmates (or in the vast majority), infection probably occurred in the seven days prior to rt-PCR; that is, between 14 and 21 days post-vaccination, before the vaccine reached maximum effectiveness. Furthermore, this effectiveness was studied in the worst possible conditions: a) in a prison outbreak, in which exposure is likely to be high; and b) with the highly transmissible SARS-CoV-2 variant B.1.1.7, which generated a high percentage of infections among vaccinated inmates. This high infection rate may well have affected the calculation of the vaccine’s effectiveness.

The infection rate was 25.9%, which is not high for an outbreak of SARS-CoV-2 B.1.1.7 in prison. This variant is between 43% and 90% more transmissible than the predecessor lineage^4^. The low rate reported is probably due to the reduction of the viral load by vaccination, as observed in studies of the AVChAdOx1 vaccine^5,6^ and the BNT162b2 mRNA Covid-19 vaccines^7^, and because the rt-PCR becomes negative more quickly in vaccinated individuals. The reduction in viral load could imply a reduction in transmission, as suggested by studies with BNT162-2 mRNA^7,8^ and AVChAdOx1 vaccine^5,6^.

Like other vaccines,^8^ the AVChAdOx1^3,5,6^ vaccine has been shown to prevent symptomatic cases. This may have contributed to the fact that in this outbreak patients were asymptomatic. The absence of symptomatic cases could be due to: a) the low viral load, as suggested above; b) the early detection of infection and the minimization of viral shedding due to the rapid isolation of inmates; and c) the drastic reduction in symptomatic cases, but not asymptomatic cases, achieved by vaccination. This reduction was also found in the study by Voysey et al^5^, in which protection with one dose in the first 90 days was 76% but protection against asymptomatic infection was only 16%. In fact, those authors doubted whether the vaccine had any impact on asymptomatic infection; they suggested that it might convert severe cases into mild cases and mild cases into asymptomatic ones, without modifying the positivity of the rt-PCR.

It is a matter of concern that 32.6% of inmates who were offered vaccination declined. Rejection was significantly higher in immigrants, who tend to have lower education and income levels, variables also associated with greater rejection by Lazarus et al^9^. Campaigns to promote vaccination are essential but may be insufficient in places such as prisons, where socio-cultural and religious ties are extremely important. In these cases, in order to increase vaccination coverage, it would be sensible to work together with ethnic, civic and religious leaders, community health agents and other respected inmates in the prison.

These results should be evaluated with care, as this is an observational study carried out at a time when the vaccine may not have reached maximum effectiveness. However, it has the advantage of feasibility, since it was carried out in the context of an outbreak in a closed institution, which made it possible to study all cases in detail.

A vaccine dose has been shown to reduce morbidity and mortality due to COVID-19^3-5,6,8^, but protection from asymptomatic infection is more controversial. In our view, two aspects are particularly important: 1. The decrease in the time to positivity of the rt-PCR, which may help to avoid transmission (always an important issue, and especially in closed facilities such as prisons) and also a shorter isolation time in some cases; 2. The fact that a single dose does little to prevent the appearance of asymptomatic cases highlights the need to maintain post-vaccination protection measures, especially if vaccination is not complete.

## Data Availability

The data that support the findings of this study are available from Prison Health Program (Catalan Institute of Health). Restrictions apply to the availability of these data as they are from the prisoners medical records, which were used under license for this study. Data are available and can be requested from Dr. Andres Marco with prior authorization from the Prison Health Program of the Catalan Health Institute.

## Acknowledgments

We thank Dr. Joan A Caylà for his extremely helpful comments and suggestions on an earlier version of the manuscript.

